# A Deep Learning Model for Screening Type 2 Diabetes from Retinal Photographs

**DOI:** 10.1101/2021.06.29.21259606

**Authors:** Jae-Seung Yun, Jaesik Kim, Sang-Hyuk Jung, Seon-Ah Cha, Seung-Hyun Ko, Yu-Bae Ahn, Hong-Hee Won, Kyung-Ah Sohn, Dokyoon Kim

**Author notes:** Kyung-Ah Sohn and Dokyoon Kim contributed equally as co-corresponding authors., **Correspondence to: Kyung-Ah Sohn**, 206, World cup-ro, Yeongtong-gu, Suwon-si, Gyeonggi-do, 16499, Republic of Korea, **Dokyoon Kim**, B304 Richards Building, 3700 Hamilton Walk, University of Pennsylvania, Philadelphia, PA, 19104-6021. Jae-Seung Yun and Jaesik Kim contributed equally as co-first authors.

## Abstract

**Objective:** We aimed to develop and evaluate a non-invasive deep learning algorithm for screening type 2 diabetes in UK Biobank participants using retinal images.

**Research Design and Methods:** The deep learning model for prediction of type 2 diabetes was trained on retinal images from 50,077 UK Biobank participants and tested on 12,185 participants. We evaluated its performance in terms of predicting traditional risk factors (TRFs) and genetic risk for diabetes. Next, we compared the performance of three models in predicting type 2 diabetes using 1) an image-only deep learning algorithm, 2) TRFs, 3) the combination of the algorithm and TRFs. Assessing net reclassification improvement (NRI) allowed quantification of the improvement afforded by adding the algorithm to the TRF model.

**Results:** When predicting TRFs with the deep learning algorithm, the areas under the curve (AUCs) obtained with the validation set for age, sex, and HbA1c status were 0.931 (0.928-0.934), 0.933 (0.929-0.936), and 0.734 (0.715-0.752), respectively. When predicting type 2 diabetes, the AUC of the composite logistic model using non-invasive TRFs was 0.810 (0.790-0.830), and that for the deep learning model using only fundus images was 0.731 (0.707-0.756). Upon addition of TRFs to the deep learning algorithm, discriminative performance was improved to 0.844 (0.826-0.861). The addition of the algorithm to the TRFs model improved risk stratification with an overall NRI of 50.8%.

**Conclusions:** Our results demonstrate that this deep learning algorithm can be a useful tool for stratifying individuals at high risk of type 2 diabetes in the general population.

## Introduction

Type 2 diabetes is one of the fastest-growing diseases in the world and poses a major threat to health globally (1). Increased prevalence of type 2 diabetes and its complications leads to increased risk of mortality in individuals with diabetes, and so adds to the already-profound social and economic burden (2). Type 2 diabetes has a long asymptomatic period before actual onset (3); however, several reports indicated that awareness of diabetes or prediabetes is not high in the general population (4,5). Therefore, regular screening for diabetes is important, particularly as the disease can be effectively prevented by selecting high-risk groups and performing appropriate interventions for prevention.

Funduscopic examination is a rapid, non-invasive, and effective tool for screening retinopathy, especially in the context of diabetes. Many studies have investigated automated screening of retinopathy using deep learning (6). Deep learning algorithms in artificial intelligence (AI) have particular strengths in analyzing image data, and a couple of studies have demonstrated equivalent or even better detection performance using deep learning compared to medical practitioners (7). It has been suggested that algorithmic disease prediction using fundus images can be applied not only to retinopathy, but also to nephropathy, neuropathy, and cardiovascular disease (8-11).

It is unclear whether a predictive algorithm comprised of a deep learning model trained on funduscopic images can detect features related to diabetes and can furthermore screen for diabetes itself. It is known that overt retinal lesions such as microaneurysms, cotton-wool spots, hard exudates, and venous beading generally appear 10-15 years after overt diabetes occurs (12). However, according to previous studies, analysis of fundus images not only captures retinal vascular or neural damage but also has considerable capacity to detect metabolic biomarkers related to type 2 diabetes such as age, blood pressure, and BMI (13). Therefore, given the accuracy of AI-based image recognition, screening for type 2 diabetes on the basis of retinal images poses a reasonable challenge for deep learning research.

The UK Biobank project is a nationwide, prospective, population-based cohort study that provides a variety of genetic, lifestyle, and clinical data, and furthermore includes over 100,000 retinal images. We primarily aimed to establish a screening model for type 2 diabetes using clinical data and funduscopic images from the UK Biobank. In addition, we aimed to investigate whether there are significant associations between the traditional risk factors (TRFs) of diabetes and funduscopic images, and also whether the diabetes risk associated with a fundus image captures either or both genetic and acquired components for diabetes. We finally tested the added value of the deep learning model and TRFs for prediction of type 2 diabetes using discrimination and reclassification methods.

## Research Design and Methods

### Study population

We used the UK Biobank dataset to develop and validate a deep learning algorithm for prediction of type 2 diabetes using retinal fundus photographs. The UK Biobank project is a prospective observational study that recruited 505,025 UK participants, aged 40-69 years at baseline, between 2006 and 2010. Each participant provided informed consent, completed a touchscreen and in-person interview with trained staff, and underwent a series of physical examinations. Extensive information was collected, including lifestyle, sociodemographic factors, medical history, biologic samples, imaging, and genome-wide genotype data. Detailed protocols for obtaining the data are available on the UK Biobank website at www.ukbiobank.ac.uk. The UK Biobank has ethical approval from the National Research Ethics Committee (June 17, 2011 [RES reference 11/NW/0382]), which was further extended (May 10, 2016 [RES reference 16/NW/0274]). Use of the UK Biobank Resource in the current study was approved under Application Number 67855. The external validation set consisted of 6,575 retinal fundus images from type 2 diabetes patients of the University-affiliated Diabetes Center of St. Vincent’s Hospital. Use of the external validation set was approved by the Catholic Medical Center Ethics Committee and conducted in accordance with the Declaration of Helsinki.

A total of 62,262 participants underwent retinal fundus imaging using a Topcon 3D OCT-1000 MKII (Topcon Corporation) during 2009-2010. All such images were obtained with a 45° primary field of view. The patients were randomly divided into three groups with no overlapping individuals: the training set, n = 37,904; the tuning set, n = 12,173; and the validation set (n = 12,185) (Supplemental Fig 1). The training and tuning sets together comprised the development set. These sets accounted for approximately 12% of patients in the UK Biobank dataset. Images of poor quality were filtered out before training and validation; only gradable retinal funduscopic images were used in this study. Detailed methods of image preprocessing for quality control are summarized in the Supplemental Materials.

**Figure 1.**
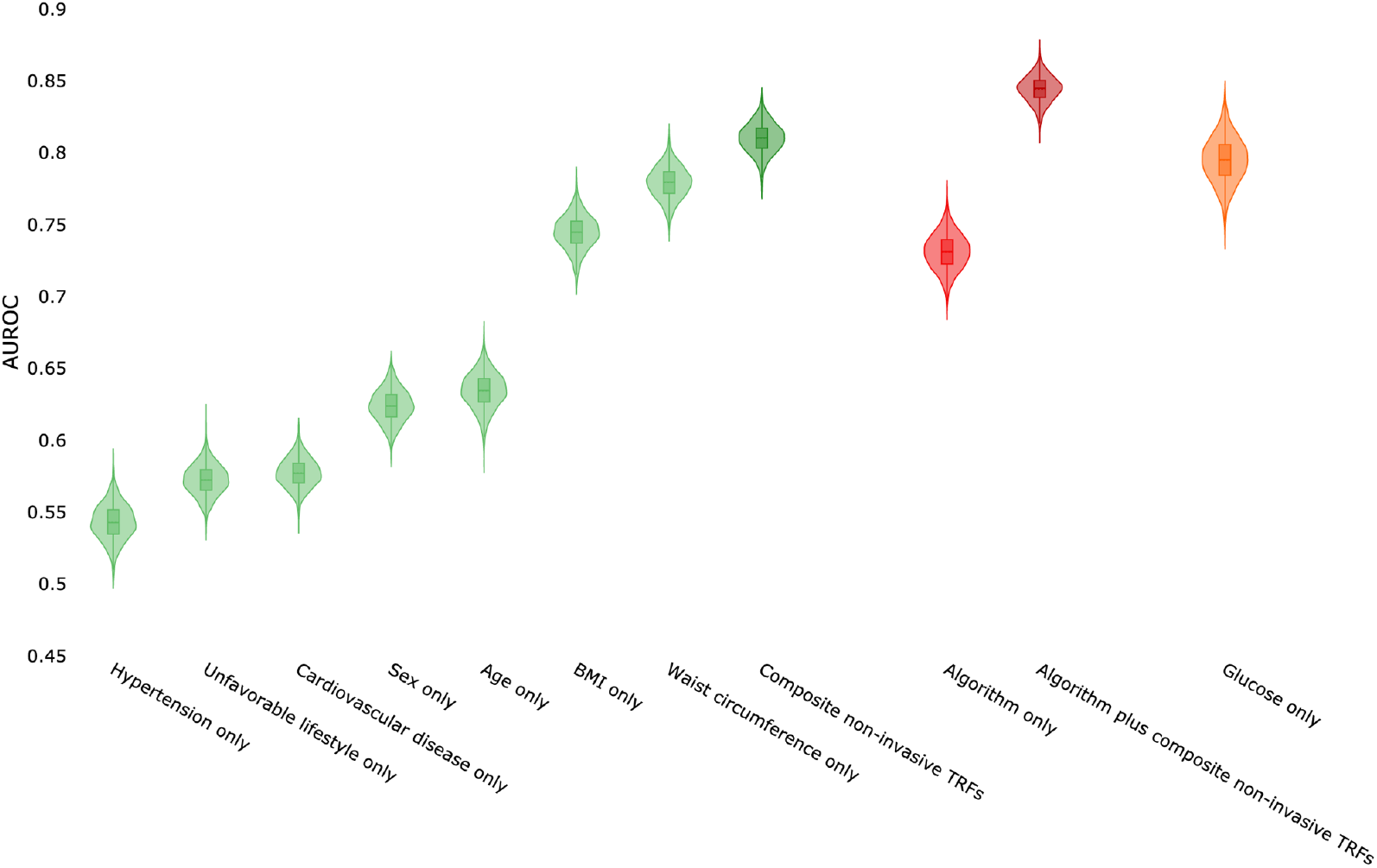
Performance comparison between each non-invasive traditional risk factor (TRF) model, composite TRFs model, deep learning algorithm, and TRFs plus algorithm model for the prediction of type 2 diabetes, determined by 2000x bootstrapping with the validation set.

### Assessment of major variables

Definition of prevalent type 2 diabetes at baseline was based on a self-report touchscreen questionnaire, nurse-led interview at enrollment, or diagnostic codes recorded across all hospital visits. We excluded subjects who reported type 1 diabetes in their verbal interview and those who had a diagnostic code for type 1 diabetes (E10). These definitions are presented in Supplemental Table S1.

Blood pressure was measured twice using the Omron HEM-7015IT digital blood pressure monitor (Omron Healthcare) or, exceptionally, a manual sphygmomanometer. We defined hypertension as systolic blood pressure ≥140 mmHg or diastolic blood pressure ≥90 mmHg. We considered four lifestyle factors: current smoking, obesity, physical activity, and dietary pattern, as recommended by the strategic goals of the American Heart Association (14). Family history of diabetes was defined as positive if either mother or father had diabetes and negative otherwise. Additional definitions and details regarding lifestyle factors and polygenic risk score for type 2 diabetes are provided in Supplemental Table S2 and Supplemental Materials, respectively.

### Algorithm development

We used ResNet18 as the deep learning algorithm, pre-trained on ImageNet data, with a resolution of 578 × 578 and taking fundus images as input (15). Since this model uses a single input image, cases where at least one eye of the patient was available were used for development. Mean squared error loss was used for the continuous target variable and cross-entropy loss for the categorical target variable during training. To prevent overfitting, we applied common augmentation methods such as flip, rotation, and crop, and implemented early stopping based on the average loss of the tuning set. Details of the model architecture and development are given in Supplemental Materials.

### Statistical analysis

For performance metrics, we used R-squared (R^2^) for continuous target variables and the area under the receiver operating characteristic curve (AUC) for binary target variables. In addition, we determined the sensitivity, specificity, positive predictive value (PPV), and negative predictive value (NPV) at an optimal threshold that maximized sensitivity plus specificity for binary classification. To evaluate the statistical significance of performance values, non-parametric bootstrapping with replacement was used to obtain metric distribution and the 95% confidence interval. We repeated 2,000 times, sampling from the validation set.

We first evaluated the capacity of the deep learning algorithm to predict TRFs associated with diabetes. Based on the literature (16), a total of eleven TRFs were selected for analysis: age, sex, hypertension, BMI, waist circumference, history of CVD, unfavorable lifestyle, triglyceride, HDL cholesterol, serum glucose, and HbA1c. Given the lack of standard criteria for determining acceptance in terms of R^2^, and assuming that some risk factors may have little effect on the retina before reaching a certain level, we also examined binary variables with discretized versions of continuous TRF variables defined on the basis of established cut-off levels for metabolic disorder: age, <60 years, ≥60 years; hypertension, <140/90 mmHg, ≥140/90 mmHg; BMI, <30 kg/m^2^, ≥30 kg/m^2^; waist circumference, <102 cm (men) or <88 cm (women), ≥102 cm (men) or ≥88 cm (women); triglyceride, <1.70 mmol/L, ≥1.70 mmol/L; HDL cholesterol, ≥1.03 mmol/L (men) or ≥1.29 mmol/L (women), <1.03 mmol/L (men) or <1.29 mmol/L (women); HbA1c (prediabetes), <5.7% or ≥5.7%; and HbA1c (diabetes), <6.5% or ≥6.5%. Next, we trained and evaluated logistic regression models for predicting type 2 diabetes using non-invasive TRFs and the deep learning algorithm. To fairly compare diagnostic performance, we determined the logistic model performances of each non-invasive TRF, the algorithm alone, and the TRFs plus the algorithm. The development set was used for logistic model learning and the validation set for performance comparison. For patients for which the algorithm made an incorrect prediction, we evaluated their characteristics as a group using cross-entropy (CE) loss. To quantify the improvement obtained by incorporating the algorithm into the TRF model, we calculated the continuous net reclassification improvement (NRI). Details concerning the calculation of NRI and CE loss are addressed in Supplemental Materials. To further assess the accuracy of our algorithm, we used follow-up data from the UK Biobank (median 8 years) to analyze the future incidence of type 2 diabetes in participants who the algorithm screened as high-risk but did not have diabetes at baseline (n = 8,889).

An additional validation test was performed using an external dataset comprised of 6,575 retinal fundus images from patients with type 2 diabetes at the University-affiliated Diabetes Center of St. Vincent’s Hospital (Supplemental Materials). To exclude the effect of overt retinal lesions in patients with diabetic retinopathy on the performance of the deep learning algorithm, we carried out sensitivity analysis after excluding participants who had diagnostic codes for diabetic retinopathy. We also performed subgroup analysis with 39,473 participants in our dataset who were eligible for both genetic analyses and funduscopic images to evaluate the association between retinal images and inherited risk for type 2 diabetes. As genetic risk markers, we used the family history of diabetes and polygenic risk score (PRS) (Supplemental Materials) (17).

All analyses were performed using Python (version 3.8.1) with the following libraries: numpy (version 1.18.5); pandas (version 1.0.5); pytorch (version 1.5.1); sklearn (version 0.23.1)

## Results

### Baseline characteristics of study population

We developed predictive algorithms for type 2 diabetes using 69,639 retinal fundus images from 37,904 patients, tuned using 22,342 images from 12,173 patients, and validated using 22,394 images from 12,185 patients, totaling 62,262 participants, all from the same UK Biobank dataset. At the time of enrollment, 7,891 (14.9%) participants had prediabetes, 2,691 (4.8%) had type 2 diabetes, and 4,149 (6.7%) had cardiovascular disease. There were no differences in clinical characteristics between the training, tuning, and validation cohorts, which were randomly divided. The demographic characteristics of the study population are shown in Table 1.

**Table 1.**
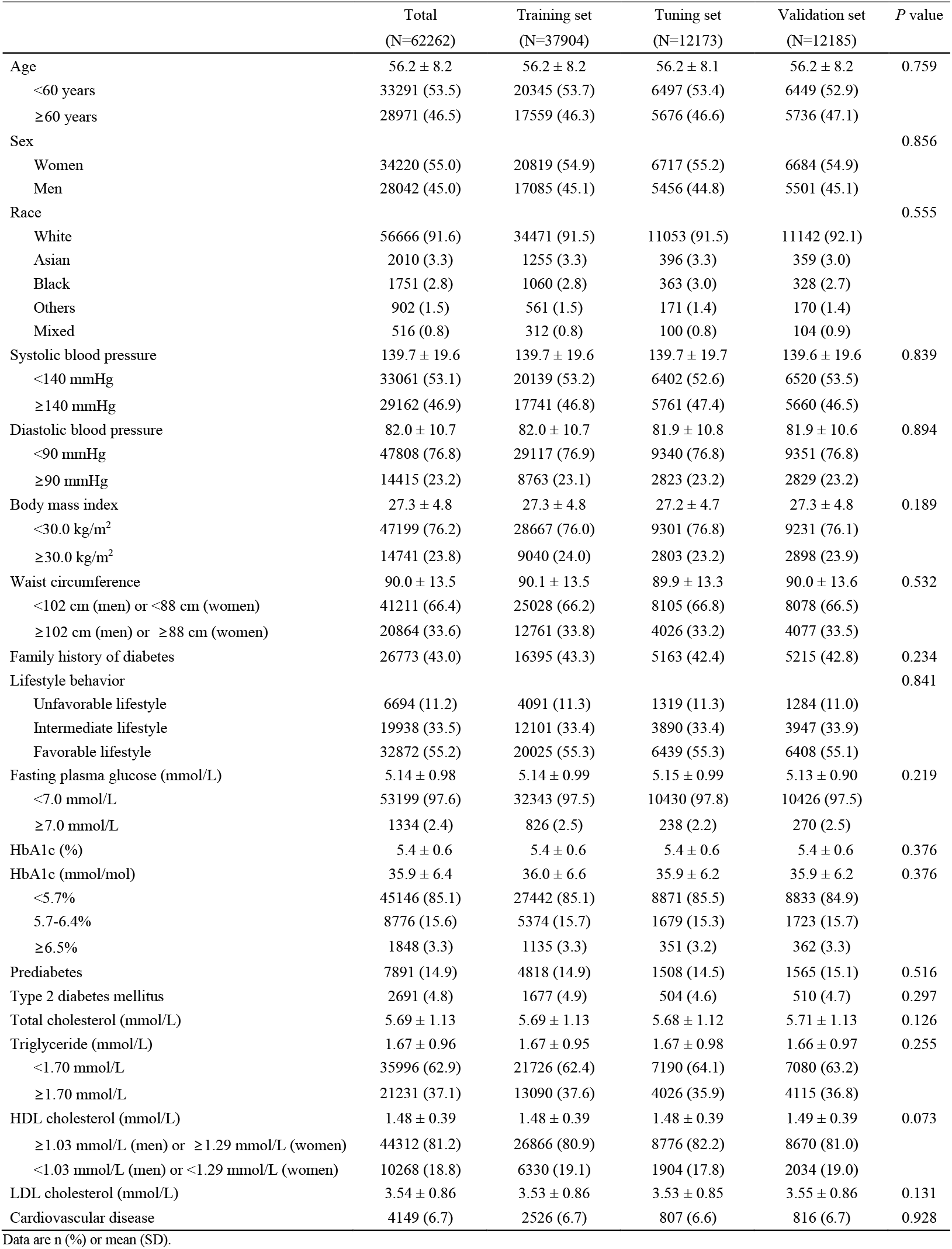
Baseline characteristics of the study population.

|  | Total<br>(N=62262) | Training set<br>(N=37904) | Tuning set<br>(N=12173) | Validation set<br>(N=12185) | *P* value |
| --- | --- | --- | --- | --- | --- |
| Age | 56.2 ± 8.2 | 56.2 ± 8.2 | 56.2 ± 8.1 | 56.2 ± 8.2 | 0.759 |
| <60 years | 33291 (53.5) | 20345 (53.7) | 6497 (53.4) | 6449 (52.9) |  |
| ≥60 years | 28971 (46.5) | 17559 (46.3) | 5676 (46.6) | 5736 (47.1) |  |
| Sex |  |  |  |  | 0.856 |
| Women | 34220 (55.0) | 20819 (54.9) | 6717 (55.2) | 6684 (54.9) |  |
| Men | 28042 (45.0) | 17085 (45.1) | 5456 (44.8) | 5501 (45.1) |  |
| Race |  |  |  |  | 0.555 |
| White | 56666 (91.6) | 34471 (91.5) | 11053 (91.5) | 11142 (92.1) |  |
| Asian | 2010 (3.3) | 1255 (3.3) | 396 (3.3) | 359 (3.0) |  |
| Black | 1751 (2.8) | 1060 (2.8) | 363 (3.0) | 328 (2.7) |  |
| Others | 902 (1.5) | 561 (1.5) | 171 (1.4) | 170 (1.4) |  |
| Mixed | 516 (0.8) | 312 (0.8) | 100 (0.8) | 104 (0.9) |  |
| Systolic blood pressure | 139.7 ± 19.6 | 139.7 ± 19.6 | 139.7 ± 19.7 | 139.6 ± 19.6 | 0.839 |
| <140 mmHg | 33061 (53.1) | 20139 (53.2) | 6402 (52.6) | 6520 (53.5) |  |
| ≥140 mmHg | 29162 (46.9) | 17741 (46.8) | 5761 (47.4) | 5660 (46.5) |  |
| Diastolic blood pressure | 82.0 ± 10.7 | 82.0 ± 10.7 | 81.9 ± 10.8 | 81.9 ± 10.6 | 0.894 |
| <90 mmHg | 47808 (76.8) | 29117 (76.9) | 9340 (76.8) | 9351 (76.8) |  |
| ≥90 mmHg | 14415 (23.2) | 8763 (23.1) | 2823 (23.2) | 2829 (23.2) |  |
| Body mass index | 27.3 ± 4.8 | 27.3 ± 4.8 | 27.2 ± 4.7 | 27.3 ± 4.8 | 0.189 |
| <30.0 kg/m² | 47199 (76.2) | 28667 (76.0) | 9301 (76.8) | 9231 (76.1) |  |
| ≥30.0 kg/m² | 14741 (23.8) | 9040 (24.0) | 2803 (23.2) | 2898 (23.9) |  |
| Waist circumference | 90.0 ± 13.5 | 90.1 ± 13.5 | 89.9 ± 13.3 | 90.0 ± 13.6 | 0.532 |
| <102 cm (men) or <88 cm (women) | 41211 (66.4) | 25028 (66.2) | 8105 (66.8) | 8078 (66.5) |  |
| ≥102 cm (men) or ≥88 cm (women) | 20864 (33.6) | 12761 (33.8) | 4026 (33.2) | 4077 (33.5) |  |
| Family history of diabetes | 26773 (43.0) | 16395 (43.3) | 5163 (42.4) | 5215 (42.8) | 0.234 |
| Lifestyle behavior |  |  |  |  | 0.841 |
| Unfavorable lifestyle | 6694 (11.2) | 4091 (11.3) | 1319 (11.3) | 1284 (11.0) |  |
| Intermediate lifestyle | 19938 (33.5) | 12101 (33.4) | 3890 (33.4) | 3947 (33.9) |  |
| Favorable lifestyle | 32872 (55.2) | 20025 (55.3) | 6439 (55.3) | 6408 (55.1) |  |
| Fasting plasma glucose (mmol/L) | 5.14 ± 0.98 | 5.14 ± 0.99 | 5.15 ± 0.99 | 5.13 ± 0.90 | 0.219 |
| <7.0 mmol/L | 53199 (97.6) | 32343 (97.5) | 10430 (97.8) | 10426 (97.5) |  |
| ≥7.0 mmol/L | 1334 (2.4) | 826 (2.5) | 238 (2.2) | 270 (2.5) |  |
| HbA1c (%) | 5.4 ± 0.6 | 5.4 ± 0.6 | 5.4 ± 0.6 | 5.4 ± 0.6 | 0.376 |
| HbA1c (mmol/mol) | 35.9 ± 6.4 | 36.0 ± 6.6 | 35.9 ± 6.2 | 35.9 ± 6.2 | 0.376 |
| <5.7% | 45146 (85.1) | 27442 (85.1) | 8871 (85.5) | 8833 (84.9) |  |
| 5.7-6.4% | 8776 (15.6) | 5374 (15.7) | 1679 (15.3) | 1723 (15.7) |  |
| ≥6.5% | 1848 (3.3) | 1135 (3.3) | 351 (3.2) | 362 (3.3) |  |
| Prediabetes | 7891 (14.9) | 4818 (14.9) | 1508 (14.5) | 1565 (15.1) | 0.516 |
| Type 2 diabetes mellitus | 2691 (4.8) | 1677 (4.9) | 504 (4.6) | 510 (4.7) | 0.297 |
| Total cholesterol (mmol/L) | 5.69 ± 1.13 | 5.69 ± 1.13 | 5.68 ± 1.12 | 5.71 ± 1.13 | 0.126 |
| Triglyceride (mmol/L) | 1.67 ± 0.96 | 1.67 ± 0.95 | 1.67 ± 0.98 | 1.66 ± 0.97 | 0.255 |
| <1.70 mmol/L | 35996 (62.9) | 21726 (62.4) | 7190 (64.1) | 7080 (63.2) |  |
| ≥1.70 mmol/L | 21231 (37.1) | 13090 (37.6) | 4026 (35.9) | 4115 (36.8) |  |
| HDL cholesterol (mmol/L) | 1.48 ± 0.39 | 1.48 ± 0.39 | 1.48 ± 0.39 | 1.49 ± 0.39 | 0.073 |
| ≥1.03 mmol/L (men) or ≥1.29 mmol/L (women) | 44312 (81.2) | 26866 (80.9) | 8776 (82.2) | 8670 (81.0) |  |
| <1.03 mmol/L (men) or <1.29 mmol/L (women) | 10268 (18.8) | 6330 (19.1) | 1904 (17.8) | 2034 (19.0) |  |
| LDL cholesterol (mmol/L) | 3.54 ± 0.86 | 3.53 ± 0.86 | 3.53 ± 0.85 | 3.55 ± 0.86 | 0.131 |
| Cardiovascular disease | 4149 (6.7) | 2526 (6.7) | 807 (6.6) | 816 (6.7) | 0.928 |

Data are n (%) or mean (SD).

### Deep learning algorithm for prediction of TRFs associated with type 2 diabetes

First, we tested and validated the capability of the algorithm to predict well-known risk factors for type 2 diabetes from fundus images. The resulting area under the curve (AUC) values obtained for the validation set are listed in Table 2. The prediction algorithm showed fair performance for HbA1c >6.5% and FBS >7.0 mmol/L, with AUC values of 0.734 (0.715-0.752) and 0.672 (0.648-0.694), respectively; however, its predictive power for triglyceride and HDL cholesterol was relatively low. Relatively high R^2^ values were obtained per one unit of age and blood pressure, and relatively low R^2^ values per one unit of BMI, waist circumference, lipid profile, glucose, and HbA1c (Table 2).

**Table 2.** Performance of deep learning algorithm for the prediction of traditional risk factors for type 2 diabetes.

| Predicted risk factor | AUC (95% CI) | Sensitivity | Specificity | PPV | NPV | R<sup>2</sup> (95% CI) per 1 unit |
| --- | --- | --- | --- | --- | --- | --- |
| Age (<60 vs. ≥60 years) | 0.931 (0.928-0.934) | 0.852 (0.847-0.856) | 0.852 (0.847-0.856) | 0.834 (0.827-0.840) | 0.868 (0.863-0.873) | 0.783 (0.778-0.789) |
| Sex (Women vs. Men) | 0.933 (0.929-0.936) | 0.854 (0.849-0.859) | 0.854 (0.849-0.859) | 0.826 (0.819-0.833) | 0.878 (0.873-0.883) | NA |
| Hypertension (BP <140/90 mmHg vs. ≥140/90 mmHg) | 0.763 (0.757-0.769) | 0.693 (0.687-0.699) | 0.693 (0.687-0.699) | 0.689 (0.681-0.697) | 0.698 (0.689-0.706) | Systolic BP 0.304 (0.294-0.314)<br>Diastolic BP 0.233 (0.223-0.244) |
| Obese (BMI <30 kg/m<sup>2</sup> vs. ≥30 kg/m<sup>2</sup>) | 0.648 (0.639-0.656) | 0.604 (0.597-0.612) | 0.604 (0.597-0.612) | 0.322 (0.312-0.332) | 0.831 (0.825-0.837) | 0.074 (0.067-0.081) |
| Central obesity (WC <102 cm (men) or <88 cm (women) vs. ≥102 cm (men) or ≥88 cm (women)) | 0.628 (0.620-0.635) | 0.590 (0.583-0.597) | 0.590 (0.583-0.597) | 0.418 (0.409-0.428) | 0.741 (0.734-0.749) | 0.144 (0.134-0.154) |
| Cardiovascular disease (no vs. yes) | 0.658 (0.644-0.673) | 0.623 (0.608-0.637) | 0.623 (0.608-0.637) | 0.105 (0.098-0.113) | 0.959 (0.955-0.962) | NA |
| Unfavorable lifestyle (no vs. yes) | 0.511 (0.498-0.523) | 0.508 (0.498-0.517) | 0.508 (0.498-0.517) | 0.894 (0.888-0.899) | 0.112 (0.107-0.118) | NA |
| Triglyceride (<1.70 mmol/L vs. ≥1.70 mmol/L) | 0.546 (0.538-0.554) | 0.534 (0.527-0.541) | 0.534 (0.527-0.541) | 0.399 (0.389-0.408) | 0.664 (0.655-0.673) | -0.028 (-0.035- -0.021) |
| HDL cholesterol (≥1.03 mmol/L (men) or ≥1.29 mmol/L (women) vs. <1.03 mmol/L (men) or <1.29 mmol/L (women)) | 0.534 (0.524-0.545) | 0.523 (0.514-0.532) | 0.523 (0.514-0.532) | 0.205 (0.197-0.214) | 0.823 (0.815-0.830) | 0.082 (0.073-0.091) |
| HbA1c (<5.7% vs. ≥5.7%) | 0.648 (0.637-0.658) | 0.600 (0.590-0.610) | 0.600 (0.590-0.610) | 0.201 (0.192-0.211) | 0.899 (0.894-0.904) | 0.061 (0.054-0.069) |
| HbA1c (<6.5% vs. ≥6.5%) | 0.734 (0.715-0.752) | 0.676 (0.656-0.693) | 0.676 (0.656-0.693) | 0.066 (0.059-0.073) | 0.984 (0.982-0.986) |  |
| Glucose (<7.0 mmol/L vs. ≥7.0 mmol/L) | 0.672 (0.648-0.694) | 0.631 (0.608-0.651) | 0.631 (0.608-0.651) | 0.043 (0.038-0.048) | 0.985 (0.983-0.987) | -0.005 (-0.009- -0.001) |

AUC, area under the curve; PPV, positive predictive value; NPV, negative predicted value; NA, not applicable; BP, blood pressure; WC, waist circumference

### Deep learning algorithm for prediction of type 2 diabetes

Next, we compared the risk factor and deep learning algorithm models in terms of diagnostic performance, and further investigated whether the deep learning model can effectively screen for the presence of type 2 diabetes based on a fundus image (Figure 1, Supplemental Table S3). In terms of performance, the composite diagnostic model using traditional non-invasive risk markers achieved an AUC of 0.810 (0.790-0.830), while that for the deep learning model using only fundus images was 0.731 (0.707-0.756). The AUC of univariate model using glucose alone was 0.795 (0.764-0.826), which is one of the diagnostic criteria for diabetes.

Among participants without type 2 diabetes, those having higher CE loss (top 20 percentile) were older, were more commonly women, were less often of European ethnicity, were more likely to have hypertension and a history of cardiovascular disease, more commonly had prediabetes, and had poorer metabolic profiles than those with lower CE loss (bottom 20 percentile). They also had higher levels of metabolic risk factors than those with lower CE loss (Supplemental Table S4). Supplementary Figure 2 shows that participants in the higher CE loss group are more similar in characteristics to the type 2 diabetes group than to those with lower CE loss (Detailed descriptions can be found in Supplemental Materials). During the median 8-year follow-up period of the UK Biobank project, type 2 diabetes developed in 134 of 8,889 participants in the validation cohort who did not have the disease at baseline. Participants with a higher CE loss tended to have higher incident rate of type 2 diabetes, with borderline significance (Supplemental Table S4, *P* for trend = 0.051).

After excluding 382 individuals with diabetic retinopathy, model performance remained essentially unaltered (AUC 0.726 [0.699-0.752]). In validation with the mixed internal and external dataset, sets with balanced labels (50% cases and 50% control) and imbalanced labels (4.7% cases and 94.3% controls) both achieved performance similar to the original results (balanced AUC, 0.703 [0.691-0.715]; imbalanced AUC, 0.703 [0.679-0.727]) (Supplemental Table S5).

We also investigated the performance of the deep learning algorithm in predicting prediabetes in our study cohort. When targeting prediabetes participants who were HbA1c ≥5.7% and <6.5% and without diabetes (n = 1723), the AUC was 0.647 (0.631-0.662) (Supplemental Table S6).

### TRFs plus deep learning algorithm model for prediction of type 2 diabetes

Adding the TRFs to the deep learning algorithm significantly improved predictive power, yielding an AUC of 0.844 (0.826-0.861). We evaluated individual classifications when adding the deep learning algorithm to the TRFs model and ultimately reclassified 3.0% of type 2 diabetes patients and 47.8% of non-diabetes patients. Overall, the addition of algorithm to the TRF model improved prediction of type 2 diabetes, with a continuous NRI of 50.8% (95% CI 40.9%-60.9%) (Table 3). Among participants who were classified in the top 10 percentile high-risk subgroup by the TRF model, overall NRI was 51.2% (95% CI 19.5-82.9).

**Table 3.** Net reclassification improvement with addition of the algorithm to traditional risk factors, determined by 2000x bootstrapping with the validation set.

| TRFs plus algorithm model |  |  |  |  | TRFs plus algorithm model |  |  |  |
| --- | --- | --- | --- | --- | --- | --- | --- | --- |
| All participants |  |  |  |  | Top 10 percentile high risk participants in TRFs model |  |  |  |
|  | Number | Individuals reclassified |  | NRI (%) | Number | Individuals reclassified |  | NRI (%) |
|  |  | Increased (%) | Decreased (%) |  |  | Increased (%) | Decreased (%) |  |
| Cases | 419 | 51.5 (46.7-56.4) | 48.5 (43.6-53.3) | 3.0 (-6.7-12.8) | 178 | 51.7 (36.1-67.4) | 48.3 (32.6-63.9) | 3.4 (-27.8-34.9) |
| Non-cases | 8889 | 73.9 (72.9-74.8) | 26.1 (25.2-27.1) | 47.8 (45.8-49.5) | 753 | 73.9 (70.8-76.8) | 26.1 (23.2-29.2) | 47.8 (41.6-53.5) |
| All | 9308 | - | - | 50.8 (40.9-60.9) | 931 | - | - | 51.2 (19.5-82.9) |

2 TRF, traditional risk factor; NRI, net reclassification improvement

### Subgroup analysis of the association between retinal images and genetic risk

To confirm whether funduscopic images can capture information concerning genetic risk for type 2 diabetes, we additionally analyzed a subgroup of 39,473 participants for whom genetic data was available after quality control (PRS, family history of diabetes). When using the deep learning algorithm for prediction of PRS, the R^2^ was low (R^2^, -0.0001 [-0.0008-0.0005]); the AUC of the algorithm for the high-genetic-risk top 20 percentile was also poor (AUC, 0.499 [0.488-0.509]) (Supplemental Table S7). Similarly, this subgroup yielded low AUC values for the prediction of type 2 diabetes when using either a PRS-based model (0.596 [0.566-0.627]) or the algorithm (0.711 [0.684-0.738]). When PRS information was added to the deep learning algorithm, predictive power was slightly improved (AUC, 0.721 [0.693-0.749]), and more so upon incorporating all genetic factors, clinical factors, and algorithms (AUC, 0.845 [0.823-0.865]) (Supplemental Table S8).

### Conclusion

The purpose of this study was to develop and validate a funduscopic image-based deep learning model for screening type 2 diabetes using the UK Biobank dataset. The main finding of our study was that the status of TRFs for type 2 diabetes, including age, sex, blood pressure, and metabolic parameters, was modestly predictable by a deep learning algorithm on the basis of retinal funduscopic images. The algorithm also showed moderate predictive performance for type 2 diabetes, which was strengthened by combining fundus imaging with TRFs; thus, incorporating fundus imaging alongside TRFs that do not require an invasive blood test can improve the discriminative power of diagnostic models for type 2 diabetes. Notably, our deep learning screening algorithm appears to capture acquired features rather than inherited features to predict type 2 diabetes.

According to major diabetes guidelines, including those from the American Diabetes Association, routine screening should be considered to identify prediabetes or type 2 diabetes in adults who have one or more risk factors, including age >45 years, family history of diabetes, history of cardiovascular disease, hypertension, dyslipidemia, and poor lifestyle habits (3). However, such screening necessitates blood sampling tests that are invasive, along with two or more visits to the clinic for medical examination, blood sampling, and confirmation of the test results. In addition, if there is no unequivocal hyperglycemic symptom, two or more abnormal results in two different samples are needed to confirm diabetes (3). On the other hand, funduscopic examination is a rapid, inexpensive, non-invasive screening tool that can be utilized even without pharmacologic pupil dilatation in an outpatient clinical setting. In previous studies, the application of a deep learning model to retinal photographs has yielded promising results in predicting systemic disease or biomarker levels. Specifically, Gerritis et al. suggested that a deep neural network can predict age, sex, blood pressure, HbA1c, and fat mass at an acceptable level (18), and Poplin et al. demonstrated that a deep learning model using only funduscopic images from the UK Biobank and EyePACS datasets achieved moderate performance at predicting six major cardiovascular risk factors, including age, sex, blood pressure, and BMI (10). Another recent publication evaluated the capacity of deep learning algorithms to predict 47 systemic biomarkers with seven Asian and European cohorts (13). These studies reported deep learning models to predict age, sex, and blood pressure well, which is in line with our results; they also reported promising performance of the models in predicting body composition indices and kidney function, measures that are associated with development of type 2 diabetes. These studies support the feasibility of funduscopic examination as a screening tool for detecting type 2 diabetes.

Several possible links are known that connect retinal structure and the risk of diabetes. For one, diabetic retinopathy shares risk factors with diabetes and its vascular complications based on the common pathophysiology; in addition, epidemiological and mechanistic evidence has consistently proposed interaction between diabetic complications (19-21). Retinal vessel tortuosity has been suggested as associated with metabolic risk factors (22), and early retinal changes such as wide retinal venular caliber have been consistently observed in prediabetes and metabolic syndrome (23). Nonetheless, to the best of our knowledge, no previous attempt has been made to predict diabetes status itself from fundus photographs using a deep learning model. Since the deep learning algorithms in our study can detect systemic biomarkers closely related to diabetes and can predict cardiovascular diseases that share common risk factors with diabetes, it is plausible to predict diabetes from fundus photographs. However, because changes in retinal structure that relate to diabetes are a long-term consequence of overt diabetes, it is challenging to determine whether deep learning can detect fundus changes related to diabetes itself. In our study, the deep learning algorithm demonstrated moderate performance in type 2 diabetes screening. Combining the deep learning model with TRFs significantly improved the discriminative power for diabetes, which means that this combined model captures additional information from retinal structure that is independent of conventional risk factors, thereby improving risk stratification. Meanwhile, we observed only a weak association of funduscopic images and genetic risk for diabetes as represented by family history or polygenic risk score. A few prior studies have considered the association between genetic determinants and retinal traits (24,25), but our results indicate that this prediction model captures features corresponding to the acquired risk of type 2 diabetes rather than the inherited risk.

We compared the performance in discrimination and risk classification of a prediction model using non-invasive TRFs and another that combines TRFs with an algorithm trained on retinal images. The new model classified nearly half of the participants more appropriately, even when only subjects in the high-risk group were analyzed. Namely, among participants who were traditionally classified as high-risk for type 2 diabetes, 47.8% of those who did not have diabetes were reclassified as lower-risk. All told, our new non-invasive predictive model incorporating a deep learning algorithm and retinal images can be used conveniently and would save time and medical costs by reducing the number of additional unnecessary blood tests or medical visits.

We further investigated per-sample CE loss to determine the characteristics of the group in which the deep learning model failed to diagnose diabetes. CE reflects the degree to which a deep learning algorithm fails to predict (26). Among participants without type 2 diabetes, those having the top 20% CE possessed more risk factors for diabetes than those in the bottom 20%. The ratio of metabolic syndrome or prediabetes in the high CE group was also significantly higher than in the low CE group. Therefore, there is a possibility that the deep learning algorithm did not fail to predict due to error; rather, this algorithm may have classified participants who are at high risk of diabetes but have not yet developed overt disease in the ‘diabetes’ group. However, our verification of the increased risk of overt diabetes in this group achieved only borderline significance, mainly due to the limited incidence of diabetes in UK Biobank follow-up data. Future longitudinal follow-up studies including larger samples are needed to clarify this hypothesis.

The results of this study should be interpreted with the following limitations. First, since the external dataset included only cases (type 2 diabetes) and we could not evaluate AUC with this dataset alone, we verified our result with a mixed internal and external validation dataset. In addition, participants in the UK Biobank are mostly European descent and middle-aged; thus, our results may not be generalizable to other ethnicities or a younger older population. However, a retinal funduscopic exam is generally used to screen for retinopathy in patients with diabetes, making it is difficult to obtain a balanced large-scale retinal funduscopic dataset that includes sufficient patients without diabetes. Moreover, very few large-scale datasets exist that include comprehensive genetic, clinical, and funduscopic image data, except for the UK Biobank dataset. Second, the diabetes prevalence among participants in the UK Biobank funduscopic image dataset was low. UK Biobank participants were recruited from the UK general population, rather than a diabetes population that needs regular screening for diabetic retinopathy (27). The prevalence of diabetes in the UK from 2009-2010, when the retinal images were collected, was reported to be approximately 5%; this is comparable with the diabetes prevalence in our study population (18). Because the prevalence of type 2 diabetes in our UK Biobank dataset was low, our algorithm exhibited high negative predictive value and low positive predictive value. There is a clear need for larger, more diverse populations in studies using retinal image recognition with deep learning models for predicting type 2 diabetes. Third, some of the information used in this study, such as lifestyle and family history, comes from participant self-reports, which may be prone to recall bias. Finally, although we tried to aggregate all available information in the UK Biobank dataset, such as questionnaires, medical records, medication history, and laboratory findings, misclassification bias may possibly have occurred.

In conclusion, we confirmed that a deep learning algorithm using only retinal fundus photographs showed considerable performance in the detection of patients with diabetes. Our results suggest a utility for funduscopic examination in combination with a deep learning model for type 2 diabetes screening in the general population. AI technology can lower the cost burden and increase the productivity of screening programs or monitoring in populations with high risk of diabetes. This is critical in light of the growing prevalence of type 2 diabetes; we expect that an AI algorithm can reduce the burden of simple screening for the disease, allowing medical practitioners to focus on more complicated aspects of care. While this technique for the detection of diabetes still needs further research and external validation in various situations, based on the results reported herein, evaluating funduscopic images with a deep learning model is a promising approach that would enable efficient screening for multiple complications related to diabetes as well as for diabetes itself.

## Data Availability

We used the UK Biobank dataset to develop and validate a deep learning algorithm for prediction of type 2 diabetes using retinal fundus photographs. The UK Biobank project is a prospective observational study that recruited 505,025 UK participants, aged 40-69 years at baseline, between 2006 and 2010. Each participant provided informed consent, completed a touchscreen and in-person interview with trained staff, and underwent a series of physical examinations. Extensive information was collected, including lifestyle, sociodemographic factors, medical history, biologic samples, imaging, and genome-wide genotype data. Detailed protocols for obtaining the data are available on the UK Biobank website at www.ukbiobank.ac.uk.

## Acknowledgments

This work was supported by the National Research Foundation of Korea Grant funded by the Korean Government (NRF-2016R1C1B1009262) and the National Research Foundation of Korea Grant (NRF-2019R1A2C1006608) funded by the Korea government. This work was also supported by NLM R01 NL012535 and NIGMS R01 GM138597.

## Author contributions

J.-S.Y. is the guarantor of this work, as such, had full access to all the data in the study and takes responsibility for the integrity of the data and the accuracy of the data analysis, and developed the study design, wrote the manuscript, analyzed and interpreted data. J.K. analyzed data, wrote the manuscript, interpreted data and contributed to discussion. S.-H.J. analyzed data, interpreted data and contributed to discussion. S.-A.C., Y.-B.A., S.-H.K., H.-H.W. revised the manuscript, contributed to discussion. K.-A.S. and D.K. interpreted results, revised the manuscript, contributed to discussion.

## Non-Standard Abbreviations and Acronyms

AI: artificial intelligence
AUC: area under the curve
CE: Cross-entropy
CVD: Cardiovascular disease
NPV: negative predictive value
PPV: positive predictive value
R^2^: R-squared
TRF: traditional risk factor

## Table legends

Supplemental Table S1. Detailed definitions of type 2 diabetes and cardiovascular disease.

Supplemental Table S2. Detailed definitions of lifestyle factors and lifestyle behavior.

Supplemental Table S3. Performance of the deep learning algorithm for prediction of traditional risk factors of type 2 diabetes.

Supplemental Table S4. Comparison of clinical characteristics according to the level of cross-entropy loss.

Supplemental Table S5. Model performance for the prediction of type 2 diabetes using internal and external validation set

Supplemental Table S6. Comparison of model performance for the prediction of type 2 diabetes and prediabetes.

Supplemental Table S7. Performance of the deep learning algorithm in predicting genetic risk for type 2 diabetes.

Supplemental Table S8. Performance of the deep learning algorithm and genetic risk plus deep learning algorithm for the prediction of type 2 diabetes

## Figure legends

Supplemental Figure S1. Summarization of the study design.

Supplemental Figure S2. UMAP visualization of image representations in the last hidden layer of the deep learning algorithm. Red colored points with 2D contour histogram represent type 2 diabetes. Blue colored points represent non-diabetes with the three groups of per-sample cross-entropy loss (High: top 20 percentile; Intermediate: 20-79th percentile; Low: bottom 20 percentile).

Supplemental Figure S3. Representative randomly-selected examples of quality assessment. *P* indicates the probability of poor quality given an image (*P*(*poor*|*image*)).

