## Supplemental Materials for "A Deep Learning Model for Screening Type 2 Diabetes from Retinal Photographs"

**Detection of invalid images**

The area of the black mask was cut such that the fundus was centered in the image. The image was squared for subsequent steps. Images for which the circular mask could not be detected were excluded in this step.

**Quality assessment**

Previous studies have reported that many of the 175,820 total UK Biobank funduscopic images are of poor quality (12% (1), 25% (2), 42% (3)). We started with a pre-trained quality assessment prediction model, the Multiple Color-space Fusion Network (MCF-Net) trained on the EyePACS dataset (http://www.eyepacs.org) of diabetic retinopathy patients provided in (4), and modified and fine-tuned the MCF-Net using some of the UK Biobank funduscopic images. This fine-tuning was necessary because of AI bias associated with ethnicity/race/origin (5). The ethnicity of patients in the EyePACS dataset definitely differs from that of patients in the UK Biobank dataset. We expect fine-tuning the MCF-Net with the UK Biobank cohort to be suitable for quality assessment. The second impetus for retraining was that MCF-Net outputs predictions of good, usable or poor quality, but in selecting a value for strict filtering of low-quality images, we needed only the two categories of good or poor quality. The UK Biobank dataset includes manual quality labeling by diabetes clinicians for 15,000 images. We split those images into a training set of 9,000, validation set of 3,000, and testing set of 3,000, then fine-tuned the model. The best performance achieved with the validation set was an area under the receiver operating characteristics (AUC) value of 0.986, and for the testing set AUC 0.983. We then applied the fine-tuned MCF-Net to the full UK Biobank dataset. Representative examples of the results are shown in Supplemental figure 3. To ensure only images with definite good quality were used in the study, we set the cut-off for a probability of poor quality at 0.05, filtering out images that scored higher.

**Model architecture**

ResNet18 is a deep learning architecture developed for recognizing natural images (6). The representative feature of ResNet is shortcut (skip) connection, which only increases the addition computation amount, has no significant impact on the number of learning parameters, and allows deep models with higher numbers of layers to learn well, even easier, and faster. According to a previous study on chest X-rays, there is no performance relationship between a model for natural images and on for chest X-ray images whether with or without pre-training (7). In addition, the performance with pre-training does not vary significantly between model types. This model is lighter than models of the Inception family, but demonstrated similar performance in analyzing chest X-rays.

**Training details**

Recent studies have reported that the pretrained parameters from natural image domain contributes to little performance improvements (7,8). Therefore, we used a ResNet18 model pre-trained on data from ImageNet. During training, the fundus images were randomly flipped, cropped, and rotated as a means of data augmentation. In our preliminary experiments, we also applied a variety of other augmentation methods, but obtained only similar or slightly decreased performance. The model was trained using an Adam optimizer, the learning rate was set to 0.0003, and the batch size was 64. For each target, the best performing model and its optimal epoch were determined by early stopping based on loss in the tuning set. All experiments were conducted in a PyTorch 1.5.1 (https://pytorch.org/) environment.

**Visualization of the model's representation of funduscopic images**

To visualize the funduscopic images as represented in the deep learning algorithm, we extracted the 512-dimensional vector representation for each image from the last hidden layer of the model and visualized them using Uniform Manifold Approximation and Projection (UMAP), a dimension reduction tool used for effective visualization (9). With regard to UMAP hyperparameters, we set the number of neighbors to 15, spread to 3, and minimum distance to 0.8. Figure 2 shows a UMAP plot of the validation set clustered into high, intermediate, and low CE loss groups, where red-colored points in the 2D contour histogram represent those with type 2 diabetes and blue-colored points represent non-diabetes samples.

**Evaluation on external set**

Since the external dataset included only cases (type 2 diabetes) and we could not evaluate AUC (the main performance metric for the internal dataset) with this dataset alone, we constructed a new dataset consisting of diabetes images from the external set and non-diabetes images from the internal validation set, which were not included in the training process. In the first experiment, we sampled a balanced set of 6,575 non-diabetes images and 6,575 diabetes images and evaluated the performance of the algorithm. In the second experiment, we constructed an unbalanced set consistent with the prevalence of type 2 diabetes (4.7%), combining 419 diabetes images with 6,575 non-diabetes images, and again determined algorithm performance. All experiments were conducted by bootstrapping 2,000 times.

**Net reclassification improvement and Cross-entropy**

In calculating the net reclassification improvement (NRI) between two prediction models, improvement was quantified as the sum of differences in the proportion of correct minus incorrect predictions for events and nonevents (Equation 1).

$$NRI = P(up \mid event) - P(down \mid event) + P(down \mid nonevent) - P(up \mid nonevent) \quad [1]$$

where upward movement ($up$) is defined as a change into a higher risk category based on the new algorithm and downward movement ($down$) as a change in the opposite direction. One of the problems with categorical NRI is that a NRI value can be changed by the categorization. Therefore, the concept of continuous NRI is more proper for comparison purposes if there are no priori criteria for categorization (Equation 2) (10).

$$\frac{1}{2} NRI(>0) = P\left(Q_{new,i} > Q_{old,i} \,\middle|\, i = event\right) - P\left(Q_{new,j} > Q_{old,j} \,\middle|\, j = nonevent\right) ] \quad [2]$$

where $NRI(>0)$ denotes "continuous NRI", $Q_{new}$ denotes the predicted probability of an event based on the "new" risk prediction algorithm, and $Q_{old}$ denotes the probability based on the "old" one. Cross entropy (CE) qualifies the difference between the predicted and true distributions (11). Per-sample CE loss (Equation 3) measures how different the predicted probability and true label are for a single sample, which can be thought as the prediction error per sample. Samples with higher CE loss are less accurately predicted, and those with lower CE loss are more accurately predicted.

$$Per\text{-}sample\ CE\ loss = -p \times log(q) - (1-p) \times log(1-q) \quad [3]$$

where $p$ denotes the true distribution of the label and $q$ denotes its predicted distribution.

**Genotyping & quality control**

UK Biobank samples were genotyped using either the Affymetrix UK BiLEVE Axiom array or the Affymetrix UK Biobank Axiom array; these have over 95% coverage and include >800,000 single nucleotide polymorphisms (SNPs). We excluded individuals having a reported sex mismatch and those with second-degree or closer relatives also in the dataset. Imputation was carried out centrally by UK Biobank researchers using the merged 1000 Genomes Project and UK 10K panels. After imputation, variant-level quality control (QC) was carried out by filtering SNPs on the basis of minor allele frequency <0.01 and imputation INFO score <0.3.

**Polygenic risk score**

To generate the type 2 diabetes mellitus polygenic risk score, we derived weights for SNPs developed by Khera et al. (12), which were determined using LDpred from genome-wide association study summary statistics from the Diabetes Genetics Replication and Meta-analysis (DIAGRAM) consortium (13). For each individual, we calculated the aggregate risk score as the sum of the risk alleles using beta coefficients for 6,630,149 SNPs with PLINK 1.90.

**Supplemental Table S1. Detailed definitions of type 2 diabetes and cardiovascular disease.**

| Disease | Path | Field ID | Code |
|---|---|---|---|
| Type 2 diabetes mellitus (at enrollment) | Verbal interview | Non-cancer illness, self-report (20002) | Diabetes (1220) |
| | | | Type 2 diabetes (1223) |
| | Touchscreen | Diabetes diagnosed by doctor (2443) | Yes |
| | First occurrence before enrollment | First reported of non-insulin-dependent diabetes mellitus (130708, 130709) | E11.x |
| | | First reported of unspecified diabetes mellitus (130714, 130715) | E14.x |
| | Medication | Treatment/medication code (20003) | Insulin (1140883066) |
| | | | Metformin (1140884600, 1141189090) |
| | | | Sulfonylurea (1141152590, 1140874744, 1140874718, 1141156984) |
| | | | Acarbose (1140868902) |
| | | | Thiazolidinedione (1141171646) |
| | | | Meglitinide (1141168660, 1141173882) |
| | HbA1c at baseline | Glycated hemoglobin (HbA1c) (30750) | |
| | Verbal interview for exclusion type 1 diabetes | Non-cancer illness, self-report (20002) | Type 1 diabetes (1222) |
| | First occurrence for exclusion type 1 diabetes | First reported of insulin-dependent diabetes mellitus (130706, 130707) | E10 |
| Type 2 diabetes mellitus (during the follow-up period) | First occurrence before enrollment | First reported of non-insulin-dependent diabetes mellitus (130708, 130709) | E11.x |
| | | First reported of unspecified diabetes mellitus (130714, 130715) | E14.x |
| | Hospital inpatient record | Hospital inpatient record, diagnosis ICD10 code (41270) | E11.x, E14.x |
| | | Hospital inpatient record, diagnosis ICD9 code (41271) | 250, 2500, 25000, 25001, 25009, 2501, 25010, 25011, 25019, 2502, 25020, 25021, 25029, 2503, 2504, 2505, 2506, 2507, 2509, 25090, 25091, 25099, 3572 |
| Diabetic retinopathy | Verbal interview | Non-cancer illness, self-report (20002) | Diabetic eye disease (1276) |
| | Touchscreen | Has a doctor told you that you have any of the following problems with your eyes? (6148) | Diabetes related eye disease |
| | Hospital inpatient record | Hospital inpatient record, diagnosis ICD10 code (41270) | H360, E113, E143 |
| | | Hospital inpatient record, diagnosis ICD9 code (41271) | 2504, 3620 |
| Coronary artery disease | Verbal interview | Non-cancer illness, self-report (20002) | 1074 (angina) |
| | | | 1075 (heart attack/myocardial infarction) |
| | Touchscreen | Vascular/heart problems diagnosed by doctor (6150) | Heart attack, angina |
| | First occurrence before enrollment | First reported of angina pectoris (131296, 131297) | I20.x, I21.x, I22.x, I23.x, I24.x, I25.x |
| | | First reported of acute myocardial infarction (131298, 131299) | |
| | | First reported of subsequent myocardial infarction (131300, 131301) | |
| | | First reported of certain current complications following acute myocardial infarction (131302, 131303) | |
| | | First reported of other acute ischemic heart diseases (131304, 131305) | |
| | | First reported of chronic ischemic heart disease (131306, 131307) | |
| Heart failure | Verbal interview | Non-cancer illness, self-report (20002) | 1076 (heart failure/pulmonary edema) |
| | | | 1079 (cardiomyopathy) |
| | First occurrence before enrollment | First reported of heart failure (131354, 131355) | I50.x |
| | Hospital inpatient record | Hospital inpatient record, diagnosis ICD10 code (41270) | I11, I110, I113, I132, I255, I420, I421, I422, I425, I428, I50, I500, I501, IO509 |
| | | Hospital inpatient record, diagnosis ICD9 code (41271) | 4254, 4280, 4281, 4289 |
| Ischemic stroke | Verbal interview | Non-cancer illness, self-report (20002) | 1081, 1082, 1583 |
| | Touchscreen | Vascular/heart problems diagnosed by doctor (6150) | Stroke |
| | First occurrence before enrollment | First reported of cerebral infarction (131296, 131297) | I63.x, I64.x |
| | | First reported of stroke, not specified as hemorrhage or infarction (131296, 131297) | |
| Peripheral artery disease | Verbal interview | Non-cancer illness, self-report (20002) | 1067 |
| | Hospital inpatient record | Hospital inpatient record, diagnosis ICD10 code (41270) | I700, I7000, I7001, I702, I7020, I7021, I708, I7080, I709, I7090, I738, I739 |
| | | Hospital inpatient record, diagnosis ICD9 code (41271) | 4400, 4402, 4438, 4439 |

| | | | |
|---|---|---|---|
| | First occurrence before enrollment | First reported of other disorders of arteries and arteriole (131390, 131391) | I77.x |
| Cardiovascular disease | Composite events of four CVD subtypes (coronary artery disease, heart failure, ischemic stroke, peripheral artery disease) | | |

CVD, cardiovascular disease

**Supplemental Table S2. Detailed definitions of lifestyle factors and lifestyle behavior.**

| Lifestyle factors | Component | Healthy lifestyle | Field ID of UK biobank |
|---|---|---|---|
| Current smoking | Current smoking at baseline | Absence | 20116 |
| Obesity | BMI at baseline | <30 $kg/m^2$ | 21001 |
| Physical activity | Number of days per week of physical activity 10+ minutes | Participating in moderate activity ≥5 days a week or vigorous activity ≥3 days a week | 884 (Moderate physical activity 10+ minutes)<br>904 (Vigorous physical activity 10+ minutes) |
| Eating habits | At least half of all following diet components was considered as a healthy lifestyle, less than half was considered as an unhealthy lifestyle | | |
| | Fruit | ≥3 serving/day | 1309 (Fresh fruit)<br>1319 (Dried fruit) |
| | Vegetable | ≥3 serving/day | 1289 (Cooked vegetables)<br>1299 (Salad or raw vegetables) |
| | Whole grains | ≥3 serving/day | 1438, 1448 (Wholemeal or wholegrain bread)<br>1458, 1468 (Bran, oat, muesli cereal) |
| | Fish | ≥2 serving/week | 1329 (Oily fish)<br>1339 (Non-oily fish) |
| | Dairy | ≥2.5 serving/week | 1408 (Cheese)<br>1418 (Milk) |
| | Refined grains | ≤1.5 serving/week | 1438, 1448 (Wholemeal or wholegrain bread)<br>1458, 1468 (Bran, oat, muesli cereal) |
| | Processed meats | ≤1 serving/week | 1349 (Processed meat)<br>3680 (Age when last ate any kind of meat, 0 if indicated having never eaten meat) |
| | Unprocessed meats | ≤1.5 serving/week | 1359 (Poultry)<br>1369 (Beef)<br>1379 (Lamb)<br>1389 (Pork)<br>3680 (Age when last ate any kind of meat, 0 if indicated having never eaten meat) |
| | Sugar-sweetened beverages | ≤1 serving/week | 6144 (Never eats sugar or foods/drinks containing sugar) |
| Lifestyle behavior | Favorable | Having at least three healthy lifestyle factors | |
| | Intermediate | Having two healthy lifestyle factors | |
| | Unfavorable | Having one or fewer healthy lifestyle factor | |

**Supplemental Table S3. Performance of deep learning algorithm and traditional risk factor models for the prediction of type 2 diabetes.**

| Risk factors used for the prediction | AUC (95% CI) | Sensitivity | Specificity | PPV | NPV |
|---|---|---|---|---|---|
| Age only | 0.634 (0.609-0.658) | 0.598 (0.565-0.629) | 0.599 (0.567-0.627) | 0.066 (0.057-0.075) | 0.969 (0.965-0.973) |
| Sex only | 0.624 (0.602-0.647) | 0.692 (0.651-0.736) | 0.555 (0.545-0.566) | 0.068 (0.061-0.076) | 0.975 (0.970-0.979) |
| Hypertension only | 0.543 (0.519-0.567) | 0.574 (0.527-0.620) | 0.512 (0.502-0.523) | 0.053 (0.046-0.059) | 0.962 (0.957-0.968) |
| BMI only | 0.745 (0.720-0.768) | 0.686 (0.665-0.711) | 0.686 (0.665-0.711) | 0.094 (0.081-0.107) | 0.979 (0.976-0.982) |
| Waist circumference only | 0.779 (0.757-0.800) | 0.705 (0.680-0.731) | 0.705 (0.686-0.725) | 0.102 (0.089-0.115) | 0.981 (0.978-0.983) |
| Unfavorable lifestyle only | 0.573 (0.552-0.593) | 0.248 (0.206-0.288) | 0.897 (0.891-0.903) | 0.102 (0.084-0.120) | 0.962 (0.958-0.966) |
| Cardiovascular disease only | 0.577 (0.559-0.597) | 0.212 (0.174-0.251) | 0.943 (0.938-0.947) | 0.149 (0.121-0.177) | 0.962 (0.958-0.966) |
| Algorithm only | 0.731 (0.707-0.756) | 0.662 (0.634-0.691) | 0.662 (0.634-0.690) | 0.085 (0.073-0.098) | 0.976 (0.973-0.980) |
| Glucose only | 0.795 (0.764-0.826) | 0.734 (0.708-0.765) | 0.734 (0.709-0.764) | 0.115 (0.100-0.135) | 0.983 (0.980-0.986) |
| Non-invasive TRFs | 0.810 (0.790-0.830) | 0.731 (0.707-0.754) | 0.731 (0.707-0.753) | 0.114 (0.098-0.129) | 0.983 (0.980-0.985) |
| Algorithm+ non-invasive TRFs | 0.844 (0.826-0.861) | 0.753 (0.732-0.774) | 0.753 (0.733-0.773) | 0.126 (0.110-0.143) | 0.985 (0.983-0.987) |

Traditional risk factors include age, sex, hypertension, obesity, central obesity, unfavorable lifestyle, and history of cardiovascular disease

AUC, area under the curve; PPV, positive predictive value; NPV, negative predicted value; TRF, traditional risk factor

**Supplemental Table S4. Comparison of clinical characteristics according to the level of cross-entropy loss.**

| | Total (N=8889) | Low cross-entropy loss (0-19th percentile) (N=1778) | Non-diabetes Intermediate cross-entropy loss (20-79th percentile) (N=5333) | High cross-entropy loss (80-99th percentile) (N=1778) | Type 2 diabetes (N=419) | *P* value |
|---|---|---|---|---|---|---|
| Age (years) | 56.1 ± 8.2 | 48.4 ± 6.9 | 56.8 ± 7.4 | 61.7 ± 5.6 | 59.9 ± 6.5 | <0.001 |
| Sex | | | | | | 0.007 |
| Women | 4938 (55.6%) | 954 (53.7%) | 2941 (55.1%) | 1043 (58.7%) | 129 (31.2) | |
| Men | 3951 (44.4%) | 824 (46.3%) | 2392 (44.9%) | 735 (41.3%) | 290 (68.8) | |
| Race | | | | | | <0.001 |
| White | 8296 (93.6%) | 1701 (95.8%) | 5026 (94.5%) | 1569 (88.6%) | 351 (84.4%) | |
| Asian | 213 (2.4%) | 30 (1.7%) | 103 (1.9%) | 80 (4.5%) | 34 (8.2%) | |
| Black | 177 (2.0%) | 16 (0.9%) | 100 (1.9%) | 61 (3.4%) | 18 (4.3%) | |
| Others | 105 (1.2%) | 10 (0.6%) | 53 (1.0%) | 42 (2.4%) | 10 (2.4%) | |
| Mixed | 71 (0.8%) | 18 (1.0%) | 35 (0.7%) | 18 (1.0%) | 3 (0.7%) | |
| Systolic blood pressure (mmHg) | 139.3 ±19.6 | 132.6 ±17.8 | 140.3 ±19.4 | 143.2 ±20.3 | 142.5 ± 17.5 | <0.001 |
| Diastolic blood pressure (mmHg) | 81.8 ±10.6 | 80.7 ±10.7 | 82.3 ±10.6 | 81.6 ±10.5 | 81.8 ± 10.1 | 0.007 |
| BMI (kg/m$^2$) | 27.1 ± 4.6 | 26.8 ± 4.5 | 27.1 ± 4.6 | 27.2 ± 4.5 | 31.6 ± 5.7 | <0.001 |
| Waist circumference (cm) | 89.3 ±12.9 | 88.0 ±12.7 | 89.5 ±13.0 | 90.1 ±12.9 | 104.0 ± 14.5 | <0.001 |
| Total cholesterol (mg/dL) | 222.1 ±42.0 | 217.6 ±40.0 | 223.1 ±42.1 | 223.8 ±43.4 | 170.6 ± 37.1 | <0.001 |
| Triglyceride (mg/dL) | 146.1 ±84.9 | 138.7 ±87.8 | 147.0 ±83.2 | 150.7 ±86.5 | 189.2 ± 120.7 | <0.001 |
| HDL cholesterol (mg/dL) | 57.9 ±14.9 | 56.7 ±13.8 | 58.1 ±15.0 | 58.9 ±15.5 | 46.6 ± 13.1 | <0.001 |
| LDL cholesterol (mg/dL) | 138.2 ±31.9 | 135.8 ±31.3 | 138.9 ±31.9 | 138.6 ±32.5 | 100.9 ± 27.2 | 0.002 |
| Fasting glucose (mg/dL) | 90.9 ±10.0 | 89.1 ± 9.2 | 91.1 ± 9.9 | 92.1 ±10.9 | 128.7 ± 52.3 | <0.001 |
| HbA1c (%) | 5.4 ± 0.3 | 5.2 ± 0.3 | 5.4 ± 0.3 | 5.5 ± 0.3 | 6.9 ± 1.2 | <0.001 |
| Prediabetes | 1286 (14.5%) | 126 (7.1%) | 754 (14.1%) | 406 (22.8%) | NA | <0.001 |
| History of CVD | 509 (5.7%) | 47 (2.6%) | 306 (5.7%) | 156 (8.8%) | 89 (21.2%) | <0.001 |
| Metabolic syndrome profiles | | | | | | |
| Metabolic syndrome (more than 3 MS components) | 2100 (23.7%) | 322 (18.1%) | 1286 (24.1%) | 492 (27.7%) | 290 (69.4%) | <0.001 |
| Abnormal waist circumference | 2817 (31.7%) | 465 (26.2%) | 1717 (32.2%) | 635 (35.7%) | 341 (67.1) | <0.001 |
| Abnormal triglyceride | 3169 (35.7%) | 563 (31.7%) | 1935 (36.3%) | 671 (37.8%) | 234 (56.0%) | <0.001 |
| Abnormal HDL cholesterol | 1506 (17.0%) | 314 (17.7%) | 885 (16.6%) | 307 (17.3%) | 187 (44.7%) | <0.001 |
| Abnormal blood pressure | 6138 (69.1%) | 993 (55.8%) | 3812 (71.5%) | 1333 (75.0%) | 329 (78.5%) | <0.001 |
| Abnormal fasting glucose level | 1192 (13.4%) | 192 (10.8%) | 717 (13.4%) | 283 (15.9%) | 281 (67.1%) | <0.001 |
| Incident type 2 diabetes during follow-up period | 134 (1.5%) | 24 (1.3%) | 72 (1.4%) | 38 (2.1%) | NA | 0.051 |

Data are n (%) or mean (SD). BP, blood pressure; CVD, cardiovascular disease; MS, metabolic syndrome

**Supplemental Table S5. Model performance for the prediction of type 2 diabetes using internal and external validation set**

| Risk factors used for the prediction | AUC (95% CI) | Sensitivity | Specificity | Positive predictive value | Negative predictive value |
|---|---|---|---|---|---|
| Algorithm using only internal validation dataset | 0.731 (0.707-0.756) | 0.662 (0.634-0.691) | 0.662 (0.634-0.690) | 0.085 (0.073-0.098) | 0.976 (0.973-0.980) |
| Algorithm using mixed 50% external set (including only type 2 diabetes) and 50% internal set (including only non-diabetes) | 0.703 (0.691-0.715) | 0.642 (0.632-0.652) | 0.642 (0.631-0.653) | 0.642 (0.631-0.652) | 0.642 (0.632-0.652) |
| Algorithm using mixed 4.7% external set (including only type 2 diabetes) and 96.3% internal set (including only non-diabetes) | 0.703 (0.679-0.727) | 0.642 (0.621-0.663) | 0.642 (0.620-0.665) | 0.078 (0.071-0.086) | 0.974 (0.972-0.977) |

AUC, area under the curve

**Supplemental Table S6. Comparison of model performance for the prediction of type 2 diabetes and prediabetes**

| Risk factors used for the prediction | AUC (95% CI) | Sensitivity | Specificity | Positive predictive value | Negative predictive value |
|---|---|---|---|---|---|
| Algorithm only for type 2 diabetes | 0.731 (0.707-0.756) | 0.662 (0.634-0.691) | 0.662 (0.634-0.690) | 0.085 (0.073-0.098) | 0.976 (0.973-0.980) |
| Algorithm only for prediabetes and diabetes (HbA1c ≥5.7% with or without type 2 diabetes) | 0.675 (0.661-0.689) | 0.621 (0.609-0.634) | 0.621 (0.609-0.634) | 0.260 (0.245-0.275) | 0.884 (0.877-0.892) |
| Algorithm only for prediabetes (HbA1c ≥5.7% without type 2 diabetes) | 0.647 (0.631-0.662) | 0.597 (0.583-0.611) | 0.597 (0.583-0.611) | 0.201 (0.187-0.215) | 0.898 (0.889-0.905) |

AUC, area under the curve

**Supplemental Table S7. Performance of the deep learning algorithm in predicting genetic risk for type 2 diabetes**

| Predicted risk factor | AUC (95% CI) | $R^2$ (95% CI) |
|---|---|---|
| Polygenic risk score (top 20 percentile high vs. bottom 80 percentile genetic risk for type 2 diabetes) | 0.499 (0.488-0.509) | NA |
| Family history of diabetes (yes) | 0.580 (0.573-0.588) | NA |
| Polygenic risk score (per 0.0001 point) | NA | -0.0001 (-0.0008-0.0005) |

AUC, area under the curve

**Supplemental Table S8. Performance of the deep learning algorithm and genetic risk plus deep learning algorithm for the prediction of type 2 diabetes**

| Risk factors used for the prediction | AUC (95% CI) | Sensitivity | Specificity | Positive predictive value | Negative predictive value |
|---|---|---|---|---|---|
| Algorithm only | 0.711 (0.684-0.738) | 0.647 (0.618-0.677) | 0.647 (0.618-0.677) | 0.073 (0.062-0.085) | 0.977 (0.973-0.981) |
| Familial history of diabetes only | 0.504 (0.476-0.533) | 0.593 (0.540-0.649) | 0.415 (0.404-0.426) | 0.042 (0.036-0.047) | 0.960 (0.953-0.967) |
| T2D PRS only | 0.596 (0.566-0.627) | 0.559 (0.532-0.586) | 0.559 (0.533-0.586) | 0.051 (0.044-0.060) | 0.967 (0.962-0.972) |
| Algorithm+familial history of diabetes | 0.711 (0.684-0.739) | 0.644 (0.617-0.677) | 0.644 (0.617-0.677) | 0.072 (0.061-0.085) | 0.977 (0.973-0.981) |
| Algorithm+T2D PRS | 0.721 (0.693-0.749) | 0.667 (0.638-0.697) | 0.667 (0.638-0.696) | 0.079 (0.067-0.093) | 0.979 (0.975-0.982) |
| Algorithm+TRFs+T2D PRS | 0.845 (0.823-0.865) | 0.762 (0.738-0.781) | 0.7620 (0.738-0.781) | 0.121 (0.104-0.138) | 0.987 (0.985-0.989) |

AUC, area under the curve; T2D PRS, polygenic risk score for type 2 diabetes; TRF, traditional risk factors for type 2 diabetes

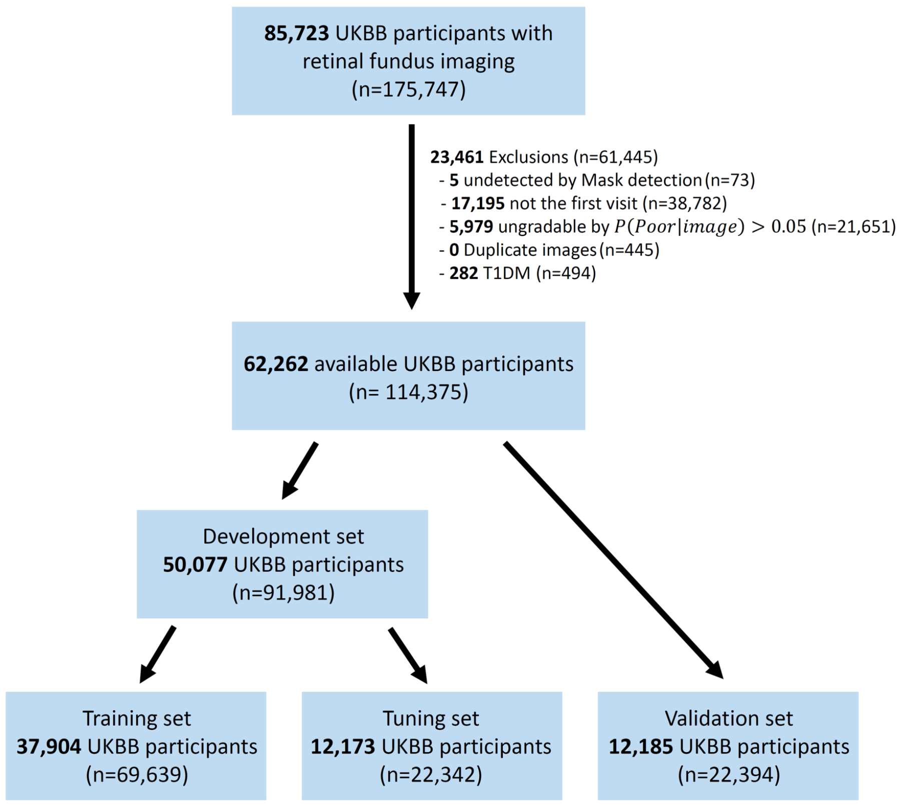


**Supplemental Figure S1. Summarization of the study design.**

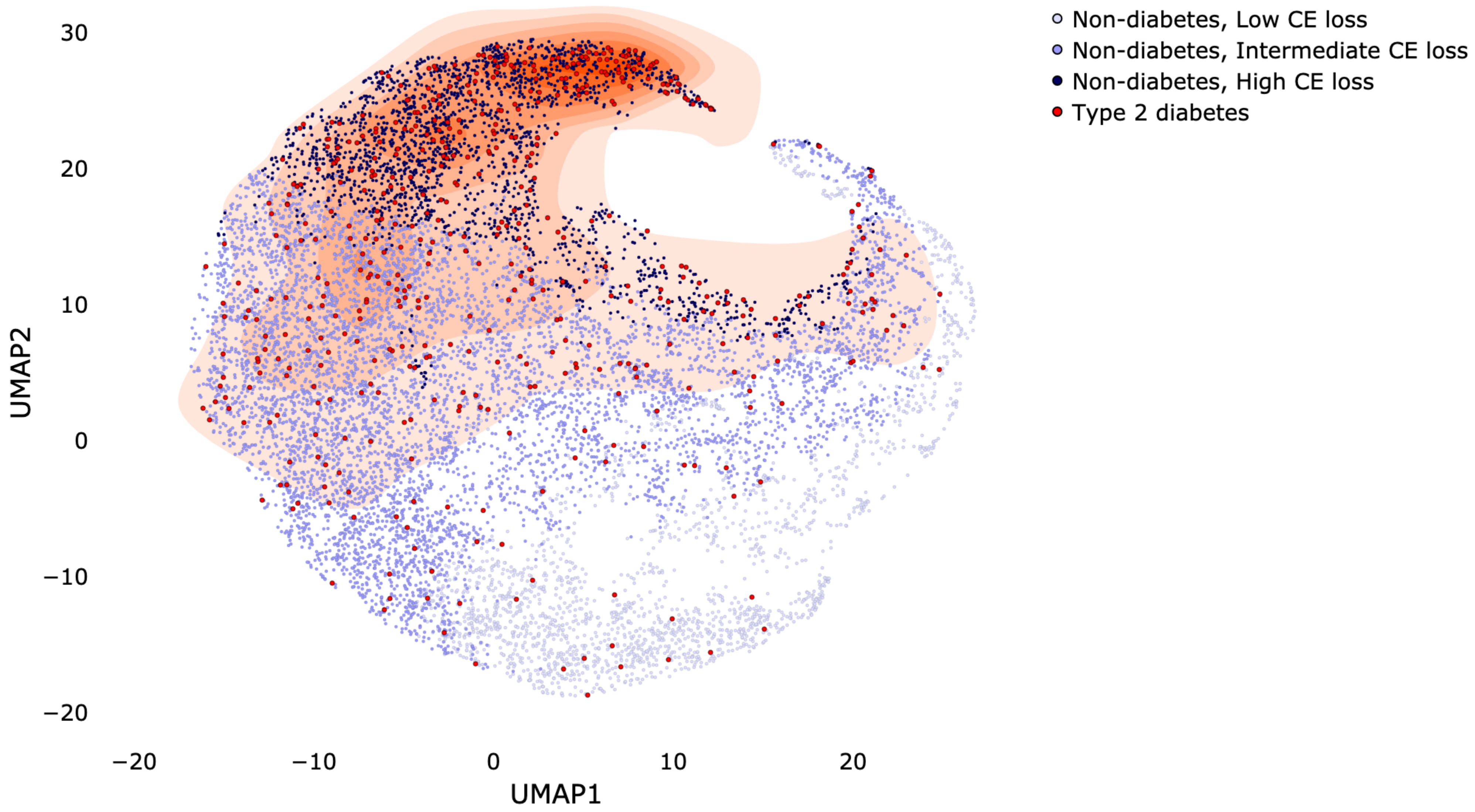


**Supplemental Figure S2. UMAP visualization of image representations in the last hidden layer of the deep learning algorithm. Red colored points with 2D contour histogram represent type 2 diabetes. Blue colored points represent non-diabetes with the three groups of per-sample cross-entropy loss (High: top 20 percentile; Intermediate: 20-79th percentile; Low: bottom 20 percentile).**

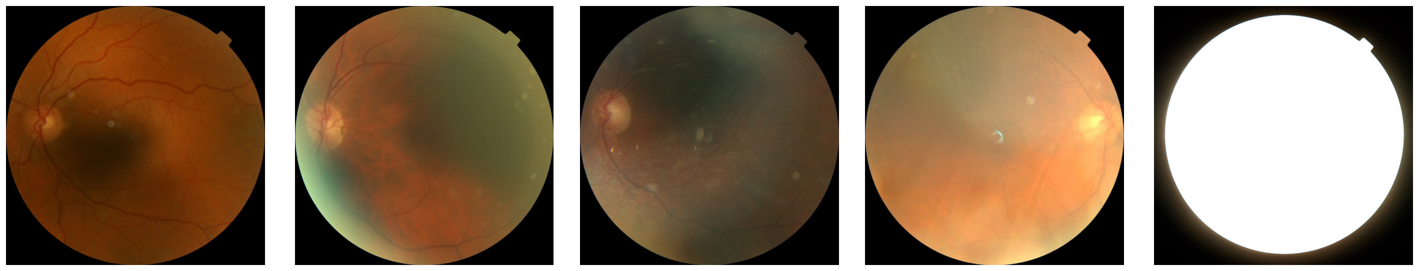

P = 0.02 P = 0.10 P = 0.30 P = 0.50 P > 0.97

**Supplemental Figure S3. Representative randomly-selected examples of quality assessment. *P* indicates the probability of poor quality given an image ($\boldsymbol{P(poor|image)}$).**

13. Scott RA, Scott LJ, Mägi R, Marullo L, Gaulton KJ, Kaakinen M, Pervjakova N, Pers TH, Johnson AD, Eicher JD, Jackson AU, Ferreira T, Lee Y, Ma C, Steinthorsdottir V, Thorleifsson G, Qi L, Van Zuydam NR, Mahajan A, Chen H, Almgren P, Voight BF, Grallert H, Müller-Nurasyid M, Ried JS, Rayner NW, Robertson N, Karssen LC, van Leeuwen EM, Willems SM, Fuchsberger C, Kwan P, Teslovich TM, Chanda P, Li M, Lu Y, Dina C, Thuillier D, Yengo L, Jiang L, Sparso T, Kestler HA, Chheda H, Eisele L, Gustafsson S, Frånberg M, Strawbridge RJ, Benediktsson R, Hreidarsson AB, Kong A, Sigurðsson G, Kerrison ND, Luan J, Liang L, Meitinger T, Roden M, Thorand B, Esko T, Mihailov E, Fox C, Liu CT, Rybin D, Isomaa B, Lyssenko V, Tuomi T, Couper DJ, Pankow JS, Grarup N, Have CT, Jørgensen ME, Jørgensen T, Linneberg A, Cornelis MC, van Dam RM, Hunter DJ, Kraft P, Sun Q, Edkins S, Owen KR, Perry JRB, Wood AR, Zeggini E, Tajes-Fernandes J, Abecasis GR, Bonnycastle LL, Chines PS, Stringham HM, Koistinen HA, Kinnunen L, Sennblad B, Mühleisen TW, Nöthen MM, Pechlivanis S, Baldassarre D, Gertow K, Humphries SE, Tremoli E, Klopp N, Meyer J, Steinbach G, Wennauer R, Eriksson JG, Männistö S, Peltonen L, Tikkanen E, Charpentier G, Eury E, Lobbens S, Gigante B, Leander K, McLeod O, Bottinger EP, Gottesman O, Ruderfer D, Blüher M, Kovacs P, Tonjes A, Maruthur NM, Scapoli C, Erbel R, Jöckel KH, Moebus S, de Faire U, Hamsten A, Stumvoll M, Deloukas P, Donnelly PJ, Frayling TM, Hattersley AT, Ripatti S, Salomaa V, Pedersen NL, Boehm BO, Bergman RN, Collins FS, Mohlke KL, Tuomilehto J, Hansen T, Pedersen O, Barroso I, Lannfelt L, Ingelsson E, Lind L, Lindgren CM, Cauchi S, Froguel P, Loos RJF, Balkau B, Boeing H, Franks PW, Barricarte Gurrea A, Palli D, van der Schouw YT, Altshuler D, Groop LC, Langenberg C, Wareham NJ, Sijbrands E, van Duijn CM, Florez JC, Meigs JB, Boerwinkle E, Gieger C, Strauch K, Metspalu A, Morris AD, Palmer CNA, Hu FB, Thorsteinsdottir U, Stefansson K, Dupuis J, Morris AP, Boehnke M, McCarthy MI, Prokopenko I. An Expanded Genome-Wide Association Study of Type 2 Diabetes in Europeans. Diabetes 2017;66:2888-2902
